# MAGNITUDE AND DETERMINANTS OF DELAYED BREASTFEEDING INITIATION AMONG MOTHERS WHO DELIVER BY CESAREAN SECTION IN A RURAL GENERAL HOSPITAL IN EAST AFRICA

**DOI:** 10.1101/2023.06.06.23290985

**Authors:** Adenike Oluwakemi Ogah, Monica Kapasa

## Abstract

**Background:** The BFHI program and timely breastfeeding initiation may be threatened by the worldwide rise in cesarean section deliveries, and this might promote prelacteal feeding. What is the current burden and determinants of delayed breastfeeding initiation among mothers, who gave birth via cesarean section?

**Subject and methods:** This was the baseline data of a prospective cohort study, where 529 healthy, singleton mother-newborn pairs were recruited consecutively at birth, from Gitwe district hospital in Rwanda.

**Results:** Overall, the burden of cesarean section delivery, delayed initiation of breastfeeding and prelacteal feeding were 38.8%, 17.0% and 6.0%, respectively. Rate of delayed breastfeeding initiation among mothers, who delivered by cesarean section was 37.6%, compared to 4.0% among those that delivered vaginally, p<0.001. Prelacteal feeds were given to 12.7% of the babies that were delivered by cesarean section, compared to 1.9% among those delivered vaginally, p<0.001. Use of probably ‘unsafe’ water sources located in the household yards was strongly linked to both cesarean section delivery (p<0.000; OR=5.71; 95%CI=2.43, 13.41) and delayed breastfeeding initiation (p<0.000; OR=44.40; 95% CI 7.97, 247.32). Mothers delivered by c-section, who were prenatally exposed to potentially harmful substances, were more likely to delay breastfeeding initiation compared to unexposed mothers (p=0.001; OR=3.14; 95% CI 1.56, 6.31). Cesarean section delivery was more likely with HIV positive mothers (p=0.010; OR=7.14; 95% CI:1.61, 33.33), teenage and entrepreneur mothers. Rate of cesarean section delivery amongst HIV positive mothers was 88.9%, compared to 36.1% among HIV negative mothers.

**Conclusion and Recommendations:** HIV positivity and non-medical causes still drive the over-use of cesarean section for delivery in this rural community, and this impacted negatively on breastfeeding initiation, thereby promoting prelacteal feeding. The uncertain safety of the water sources and prenatal exposures to harmful social habits also need to be addressed.

## Background

Breastfeeding and baby friendly hospital initiative (BFHI) programs are some of the WHO primary health care strategic packages aimed to secure infant survival. The benefits of breastfeeding to the mother, infant and environment have been well documented and these benefits act in a dose-response relationship, whereby increasing breastfeeding duration results in more infant health benefits.^1–3^ Timely initiation of breastfeeding is one of the most effective ways to ensure newborn survival and wellbeing.^4^ WHO recommends that breastfeeding should be commenced within 1 hour after birth regardless of mode of delivery. C-section delivery, in practice, however, may not allow breastfeeding initiation up to 4-6 hours after birth. Delays in breastfeeding initiation accompanying c-section delivery are associated with maternal/infant separation (lack of skin-to-skin contact), reduced suckling ability, decreased infant receptivity, and insufficient milk supply, which are predictive of shortened breastfeeding duration.^5–8^ Skin-to-skin contact has been suggested to improve breastfeeding initiation, maintenance and duration.^9^ Timely initiation of breastfeeding, amongst other benefits reduces the risk of late onset sepsis, necrotizing enterocolitis (NEC), sudden infant death syndrome (SIDS) and improves neonatal mortality, especially in resource limited countries^10–12^. Breastfeeding within the first hour post-delivery has also been cited as an important predictor of continued breastfeeding^13–15^ as a result of higher levels of prolactin release in these mothers, compared to those, who delayed in initiating breastfeeding.^16–20^ Oxytocin-related lowered stress levels in mothers, who promptly initiated breastfeeding, have also been noted to sustain continued breastfeeding.^21^

According to WHO global report [12], only 39.7% (60.3% delayed) of women, who had c-section delivery started to breastfeed their infant within one hour of birth [22, 23]; rural residence and poor socioeconomic status, rather than urban residence and wealth, were protective factors for timely breastfeeding initiation and avoidance of prelacteal feeds. Socioeconomic factors [24, 25] have been shown to contribute significantly to the increase in c-section use in low-middle income countries (LMICs).

The burden of timely breastfeeding initiation is lower in Ethiopia (44-76%) compared to Rwanda (84% at national level and 86% in the rural areas).^26, 27^ Burundi, a sister country in East Africa has timely breastfeeding initiation rate, similar to that of Rwanda, at 86.19%. ^28^ In the 2020 RDHS report,^29^ overall, 4% of the babies (3% in the rural areas), whose first breastfeed were delayed, received prelacteal feeds in Rwanda. The current status of breastfeeding initiation among mothers, who delivered by c-section in Rwanda and in other sub-Saharan African (sSA) countries is not known and is likely to higher than the rate in the general population.

Cesarean section (c-section) is an important medical intervention for reducing the risk of poor perinatal outcomes. WHO recommends that cesarean section be done only when medically indicated. HIV positivity is not an indication for cesarean section delivery, according to WHO. Medical indications for c-section include placenta previa, breech presentation, contracted pelvis and post-term pregnancies^8, 30, 31^. Non-medical factors leading to c-section, include maternal age, socioeconomic status, literacy level, occupation, religion and culture^32–34^. C-section rates above 10% and for non-medical reasons indicate overuse and are counterproductive.^35, 36^ C-section rates between 10% and 15% are linked with a decrease in maternal and neonatal mortality. C-section rates under 10% may indicate unmet requirements.

In developed places, such as Canada, c-section rate was 27.1% as at 2012^32, 37^ and it has been observed to adversely affect breastmilk initiation and duration.^13, 38^ Sub-Saharan African (sSA) nations have experienced rising trend of c-section deliveries, with wide variations (ranging from 1% in South Sudan to 16% in Ghana as of 2018); and high rates of preventable maternal and neonatal mortalities.^39–41^ As at 2021, the lowest rate of c-sections deliveries in sub-Saharan Africa had risen to 7.3% .^42^ Uganda, a close neighboring country, has a national c-section delivery incidence of 6%.^43^

C-section rates have likewise steadily increased over time in Rwanda - from 2% in 1992 and 2000 to 3% in 2005, 7% in 2010, 13% in 2014–15, 15.6% in 2019–20, and now 25.9% in 2021 ^43–46^ – as well as a notable decline in maternal and child mortality between 2000 and 2017, unlike in other sub-Sahara African nations [39]. In Rwanda, the incidence of c-sections is roughly four times higher in private (60.6%) than in public (15.4%) health facilities; highest in Kigali capital city (26.4%) and in the southern province (16.2%), where the current study site is located. The lowest rate of c-section delivery was found in the eastern province of Rwanda, at 11.2%, showing large disparities across the provinces in the same country ^30, 36^.

Despite the increasing use of c-section for deliveries, neonatal morbidity and mortality rates continue to be alarmingly high in Rwanda, particularly in rural areas, suggesting that the increase in c-section rates has yielded no added benefits for the neonates. ^45, 46^ Therefore, the rising trend in the cesarean section delivery rates, may be a barrier to the continued success of the WHO-BFHI program, which was created to improve newborn survival.

The effects of c-section delivery on breastfeeding initiation have not been extensively studied in resource-limited settings, especially in the rural areas. Data from Gitwe Hospital/clinic (study site) have not featured in any of the RDHS survey publications. Sharing Kibe et al.’s ^30^ view, there is need to examine the high regional and within country disparities in the c-section trends, that exist within sSA nations, and its impact on breastfeeding initiation. Understanding the predictors of delayed initiation of breastfeeding could help identify modifiable risk factors and facilitate improvement in breastfededing initiation practices. This current study, that took place in a poorly researched remote community in Rwanda, contributed to this knowledge and will assist decision and policy makers to develop tailored guidelines and interventions, to ensure proper use of c-section for deliveries, further strengthen the BFHI program and improve newborn survival. Hence, the objectives of this study, were to determine the magnitude of and investigate the relationships between delayed breastfeeding initiation, prelacteal feeding and c-section delivery in a rural general hospital and clinic in Rwanda.

## Concept of the study

Figure 1, shows the maternal, neonatal and household determinants of cesarean section delivery, that were investigated in this study. In turn, the effect of cesarean section delivery on delayed breastfeeding initiation and promoting prelacteal feeding as a secondary outcome, were also studied.

**Fig. 1:**
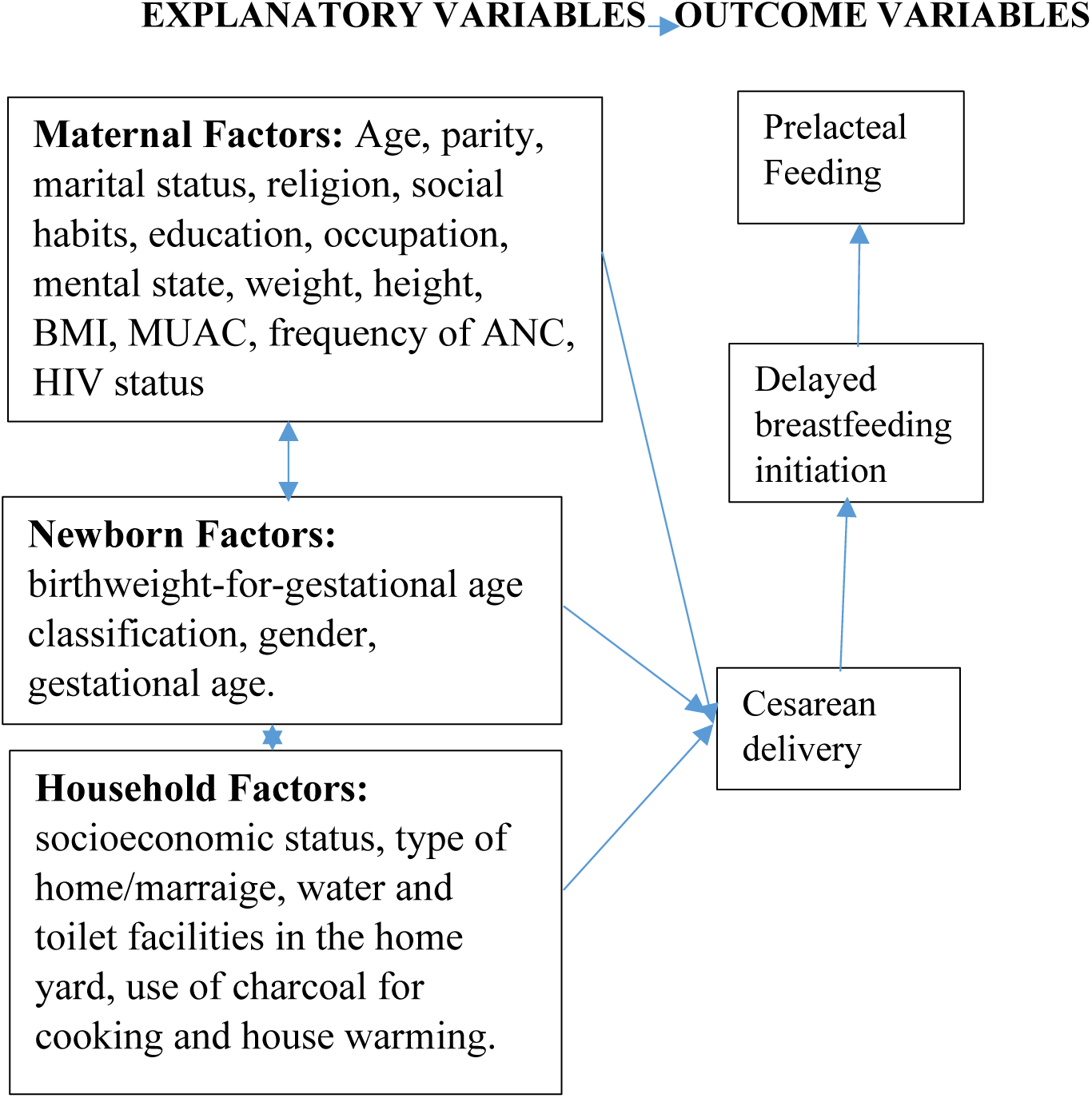
Concept of the study

## Materials and methods

The methods employed in carrying out this study is discussed in this section.

### Study setting

Rwanda is a low-income, agricultural and landlocked country with approximately 11 million people living in five provinces, covering an area of 26,338 km^2^. It is called the home of a ‘1000 hills’^44–46^. The limited area of flat land available in most part of Rwanda is a hindrance to farming, animal rearing and construction of standard residential houses, among others. For example, the recommended minimum of 50 feet distance between source of household water and sewage tank in residential yards are often compromised during construction, leading to contamination of water source. There are 2 peak raining seasons in the country: April to June and September to November.

Rwanda has an average of 4.4 persons per household ^46^ and a gross domestic product per capita of US $780.80.^46^ About half (48%) of its population is under 19 years of age and 39% live below the poverty line, with a life expectancy at birth of 71.1 years for women and an adult literacy rate of 80% among 15–49 years old women. In addition, 87.3% of the population have health insurance and access to health services; spending an average of 47.4 min to reach a health centre.^46^

Non-availability of regular supplies of clean and safe piped water, has been a longstanding problem in Rwanda, as a whole, probably because of its landlocked and hilly terrains, making construction and supply of piped water, a major challenge. Because of the infrequent flow of piped water, the taps and pipes may be rusted and breached in some places, especially in the rural areas, further leading to contamination of household water. Many families store rain water in big tanks for use in their homes and this may become polluted (in the writer’s opinion) because of difficulties of cleaning these storage tanks. A few non-profit organizations, such as USAID and Water-for-life have sunk bore holes in strategic locations in a few number of villages (including Gitwe) in the country, with the aim of alleviating this water problem.^45, 46^

Gitwe village is located on a high altitude of 1,674 meters above sea level, in the southern province, 240km from Kigali, which is the capital city of Rwanda. Gitwe District hospital began in 1995, immediately after the genocide, for the purposes of providing medical services and later training to this isolated community. The hospital currently has 100% government support, since year 2020. The maximum number of deliveries at the hospital per month was about 200. Some of the challenges in the hospital include poor specialist coverage and few trained health workers, poor supply of equipment, water, electricity, laboratory services and medicines. Challenging cases are referred to the university of Rwanda teaching hospitals in Butare or Kigali. Gitwe village was selected for this study, because there was no published birth data from this poorly researched and remote community. In 2019, birth, feeding and growth data on 529 healthy mother-singleton newborn pairs were compiled in this village, over a period of 12 months for this study, which was carried out in the delivery room and postnatal ward of Gitwe District hospital and at its annex, the maternal and child health clinic.

### Data source and sample

This was a prospective cohort study design. Mother-newborn pairs were recruited consecutively, on first-come-first-serve basis. Maternal file review and newborn anthropometry [weight (kg), length (cm) and head circumference (cm) measurements, recorded to the nearest decimals] were carried out, soon after birth. Newborn gestational age was determined using the maternal last menstrual period (LMP) and expanded new Ballard criteria. Newborn classification into small-for-gestational age (SGA), appropraite-for-gestational age (AGA) and large-for-gesttaional age (LGA) was based on weight and gestational age. Socio-demographic and neonatal feeding (including prelacteal feeding) data were obtained. Time intervals from birth to breastfeeding initiation were recorded in minutes and hours at the postnatal ward. Delayed breastfeeding initiation was defined as breastfeeding initiation interval beyond 1 hour from birth, regardless of the mode of delivery. Mothers were interviewed using semi-structured questionnaires and measured at 6 weeks post-partum in the postnatal clinic. The questionnaires were read to the mothers and filled by the research assistants.

To ensure the quality of data collected, 2 registered nurses were trained as research assistants at Gitwe Hospital for 2 days on the over-all procedure of mother and newborn anthropometry and data collection by the investigator. The questionnaires were pre-tested before the actual data collection period, on 10 mother-infant pair participants (2% of the total sample). The investigator closely followed the day-to-day data collection process and ensured completeness and consistency of the questionnaires administered each day, before data entry.

### Statistical analysis

Data clean up, cross-checking and coding were done before analysis. These data were entered into Microsoft Excel statistical software for storage and then exported to SPSS version-26 for further analysis. Both descriptive and analytical statistical procedures were utilized. Participants’ categorical characteristics were summarized in frequencies and percentages. Numerical characteristics were presented in means and standard deviation for normally distributed data; and median and interquartile ranges for skewed data. Factors determining mode of birth (c-section or vaginal) and the interaction between c-section and breastfeeding initiation (delayed or timely) were investigated for significance using Mann Whitney U (for numerical data); cross-tabulations/chi-square tests (for categorical data), as appropriate. Logistic regression models (binary logistic and multivariate linear) were created to examine the relationships between the independent variables (maternal, neonatal and household characteristics) and dependent variables (mode of delivery and breastfeeding initiation), controlling for confounding and to generate the odds ratio. Factors with p-values <0.1 were included in the regression model. Odds ratio (OR), with a 95% confidence interval (CI) were computed to assess the strength of association between independent and dependent variables. For all, statistical significance was declared at p-value < 0.05. The reporting in this study were guided by the STROBE guidelines for observational studies.^47^.

### Ethics

Ethical approval from the Health Sciences Research Ethics Committee of the University of the Free State in South Africa (Ethical Clearance Number: UFS-HSD2018/1493/2901) was obtained. Written permission to collect data was obtained from the Director of Gitwe District Hospital and the eligible mothers gave their informed consent before enrolment. The participants were given research identity numbers and the principal investigator was responsible for the safe keeping of the completed questionnaires and collected data, to ensure anonymity and confidentiality of the participants.

### Results

The following are the results obtained from the study.

### Participants

Five hundred and ninety-seven (597) babies were delivered at Gitwe Hospital, Rwanda, between 3^rd^ January and 9^th^ May 2019, out of which, eligible 529 mother-newborn pairs were enrolled into the study, Figure 2.

**Fig 2:**
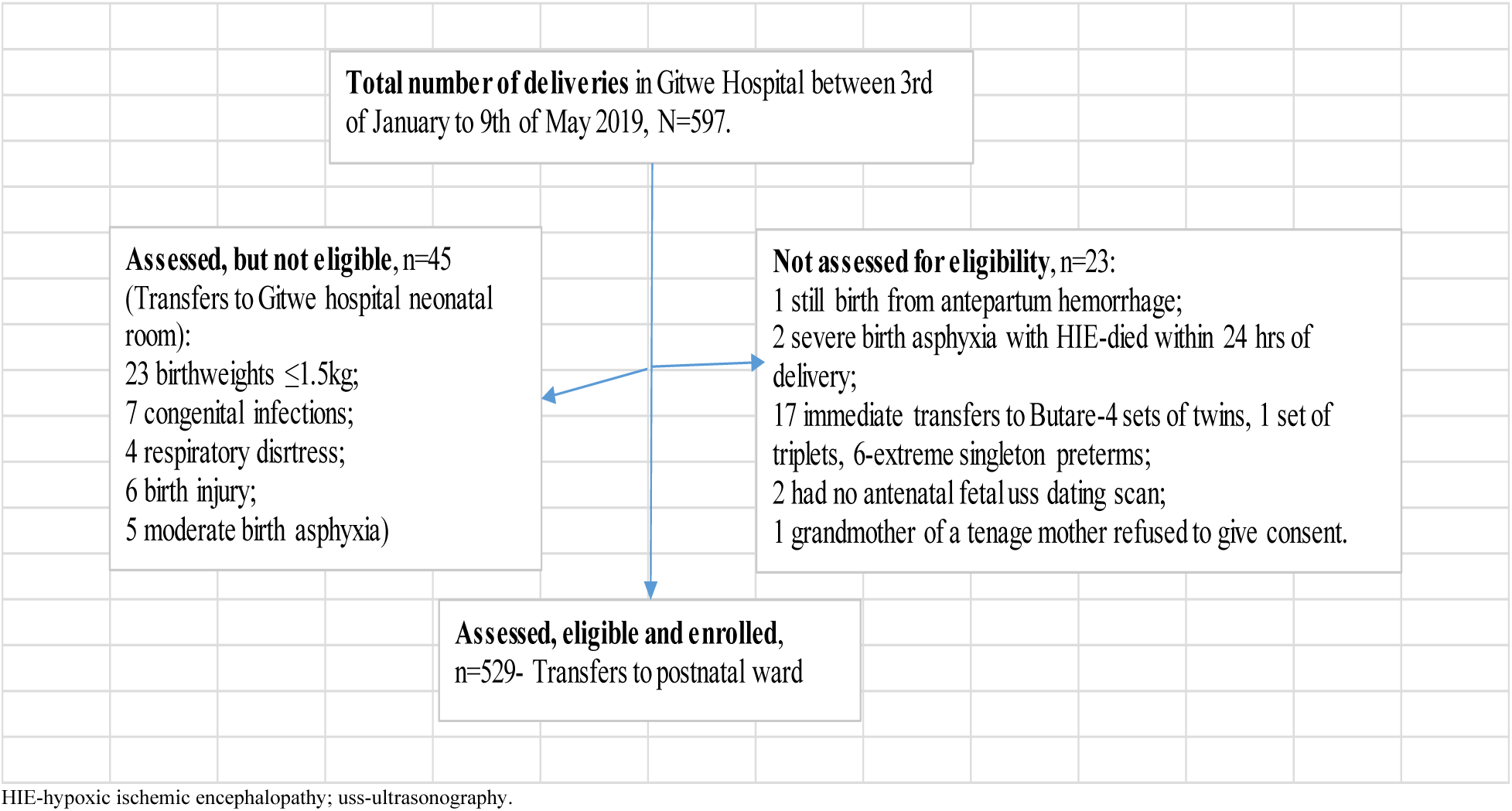
Flow of participants from admission to recruitment into study

Percentages of small for gestational age (SGA), appropriate for gestational age (AGA) and large for gestational age (LGA), were 21.4%, 71.6% and 7.0%, respectively, Table 1. The majority were male (53.5%) and term babies (57.5%).

**Table 1:**
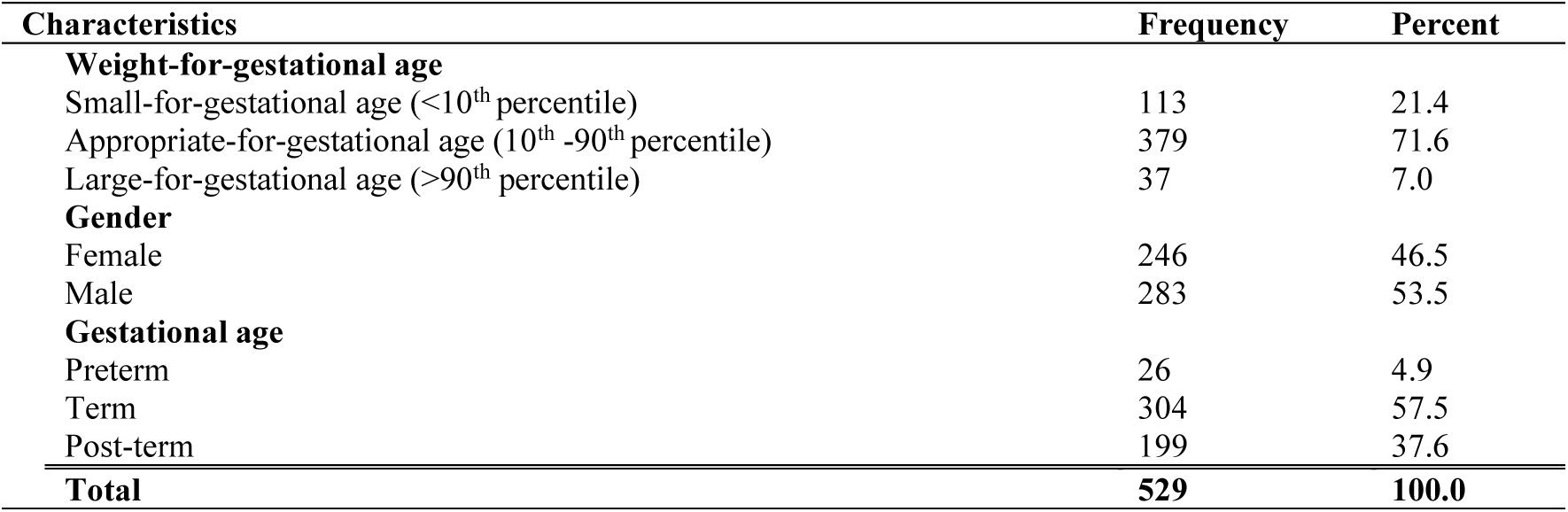
Newborn characteristics in the study

The median age, parity, duration of schooling, weight, height and BMI of the mothers were 28 years, 2.0, 6 years, 62.0kg, 159cm and 24.7kg/m^2^, respectively. The highest percentage of the mothers in the study, were young in age (43.9%), of primary school education (65.8%), unemployed/unskilled (58.8%), married (90.7%), christians (97.5%), middle class (75.0%), moderate parity (51.4%). Three (0.6%) mothers did not attend antenatal care clinic, and the majority (56.9%) had insufficient ANC visits (1-3). HIV positivity rate amongst the mothers was 5.1%, and 17.0% (90 out of 529) of the mothers delayed in commencing breastfeeding. Cesarean section delivery and prelacteal feeding rates were 38.8% and 6.0% respectively. Two babies (0.4%) were not delivered in any health facility, Table 2.

**Table 2:** Categorical characteristics of mothers and their households in the study (n=529)

| **Characteristics** | **Frequency** | **Percent** |
| --- | --- | --- |
| **Age (completed years)** |  |  |
| ≤18 (Teenage mother) | 23 | 4.3 |
| 19-25 (Very young mother) | 195 | 36.9 |
| 26-35 (Young mother) | 232 | 43.9 |
| >35 (Elderly mother) | 79 | 14.9 |
| **Education** |  |  |
| None/Informal | 26 | 4.9 |
| Primary | 348 | 65.8 |
| 'O' level secondary | 105 | 19.8 |
| 'A' level secondary/College | 32 | 6.0 |
| University | 18 | 3.4 |
| **HIV status** |  |  |
| Positive | 27 | 5.1 |
| Negative | 502 | 94.9 |
| **Occupation** |  |  |
| Unemployed/Unskilled | 311 | 58.8 |
| Semi-skilled/Skilled technician | 195 | 36.9 |
| Shop owner | 13 | 2.5 |
| Professional | 10 | 1.9 |
| **Marital status** |  |  |
| Not married (single/separated) | 49 | 9.3 |
| Married | 480 | 90.7 |
| **Religion** |  |  |
| Islam | 13 | 2.5 |
| Christian | 516 | 97.5 |
| **MUAC (cm)** |  |  |
| Low (<23) | 25 | 4.7 |
| Not low (≥23) | 504 | 95.3 |
| **BMI (kg/m<sup>2</sup>)** |  |  |
| Thin (<18.5) | 4 | 0.8 |
| Not thin (≥18.5) | 525 | 99.2 |
| **Maternal height (cm)** |  |  |
| Short (<150) | 39 | 7.4 |
| Not short (≥150) | 490 | 92.6 |
| **Parity** |  |  |
| Primiparous (1) | 196 | 37.1 |
| Moderate parity (2-4) | 272 | 51.4 |
| Multiparous (≥5) | 61 | 11.5 |
| **Mode of delivery** |  |  |
| Cesarean section | 205 | 38.8 |
| Vaginal delivery | 324 | 61.2 |
| **Place of delivery** |  |  |
| Outside health facility | 2 | 0.4 |
| Inside health facility | 527 | 99.6 |
| **Time of commencement of first breastfeed** |  |  |
| Delayed | 90 | 17.0 |
| In time | 439 | 83.0 |
| **Prelacteal feeding** |  |  |
| Yes | 32 | 6.0 |
| No | 497 | 94.0 |
| **Socioeconomic status** |  |  |
| Destitute/Poor | 115 | 21.8 |
| Middle | 397 | 75.0 |
| Rich | 17 | 3.2 |
| **Exposure to potentially harmful social habits** |  |  |
| Yes | 66 | 12.5 |
| No | 463 | 87.5 |
| **Availability of water source in household yard** |  |  |
| No | 355 | 67.1 |
| Yes | 174 | 32.9 |
| **Availability of toilet facility in household yard** |  |  |
| No | 313 | 59.2 |
| Yes | 216 | 40.8 |
| **Use of charcoal for cooking and house warming** |  |  |
| Yes | 509 | 96.2 |
| No | 20 | 3.8 |
| **Frequency of visits for antenatal care** |  |  |
| 0 to <4 | 301 | 56.9 |
| $\geq 4$ | 228 | 43.1 |
| **Total** | **529** | **100.0** |

Fifty-one (77.3%) out of the 66 mothers that were exposed to potentially harmful habits during pregnancy, ingested herbs (content of herbs were unknown), 9 (13.6%) ingested alcohol (amount unknown), 13 (19.7%) were exposed to tobacco smokes from relatives living in the same house. Five (7.6%) were exposed to both herbs and alcohol, while 2 (3.0%) were exposed to both herbs and tobacco. Majority of the mothers had good BMI (99.2%), use charcoal as fuel (96.2%), did not have water (67.1%) nor toilet (59.2%) facilities in their home yards, Table 2.

### Chi test of factors significantly associated with mode of delivery

Two hundred and five (38.8%) of the 529 mothers delivered by c-section in this study. Independently, the following factors were significantly associated with cesarean section delivery after chi square test: teenage motherhood (p=0.092, 13, 56.5%), maternal obesity (p=0.005, 17, 68.0%), short mothers (p<0.000, 29, 74.4%), HIV positive mothers (p<0.000, 24, 88.9%), maternal entrepreneur occupation (p<0.000, 10, 76.9%), maternal high parity (p=0.503, 26, 42.6%), high (>6) number of antenatal clinic visits (p<0.000, 8, 80%), delay in initiating breastfeeding (p<0.000, 77, 85.6%), prelacteal feeding (p<0.000, 26, 81.3%), exposure to potentially harmful social habits (p<0.000, 42, 63.6%), use of water facilities located in household yards (p<0.000, 128, 73.6%) and LGA birth (p=0.001, 25, 67.6%). Though, not statistically significant, the rich and highly educated mothers were also more likely to deliver by c-section than their poor or middle-class (52.9% vs 38.3%) and primary school leaver (66.7% vs 31.0%) counterparts, respectively.

### Binary logistic regression analysis of factors determining mode of delivery

Seven factors remained significantly associated with cesarean section delivery after regression analysis:-maternal HIV status, parity, maternal age, use of charcoal for cooking and house warming, occupation, time of commencing first breastfeed and use of household yard water facility.

Delivery by cesarean section was less likely associated with mothers, who were HIV negative (p=0.003; OR=0.11; 95% CI= 0.03, 0.48), that commenced their first breastfeed within 1 hour after birth (p<0.000; OR=0.18; 95% CI=0.08, 0.41), that were of moderate parity (p=0.031, OR=0.31; 95% CI=0.11, 0.90), do not use charcoal for cooking and house warming (p=0.037, OR=0.23; 95% CI=0.06, 0.91). However, delivery by cesarean section was more likely associated with mothers, who were young (p=0.046; OR=2.54; 95% CI=1.02, 6.36) and elderly (p=0.034; OR=2.18; 95% CI=1.06, 4.49), who were engaged in semi-skilled or skilled labor (p=0.027; OR=6.74; 95% CI=1.24, 36.60); entrepreneurs and professionals in their occupation (p=0.018; OR=7.44; 95% CI=1.41, 39.41); and who utilize the water facilities located in their household yards (p<0.000; OR=5.71; 95%CI=2.43, 13.41), compared to their counterparts.

### Multivariate linear regression analysis of factors associated with time of breastfeeding initiation among babies delivered by cesarean section

The average time interval from birth to breastfeeding initiation was 0.18hours shorter among mothers, who were not exposed to potentially harmful habits, compared to those who were exposed prenatally (p=0.013). The average time interval from birth to breastfeeding initiation was 0.18hours shorter among married mothers compared to that of single mothers (p=0.015), Table 3. Although, gender of the baby did not show a significant relationship (p=0.085) with time interval to breastfeeding initiation, gender was the strongest factor in this study, determining time interval to breastfeeding initiation (B=0.12). The average time-interval from birth to breastfeeding initiation was 0.12hours shorter among the female newborns compared to that of the male newborns.

**Table 3:**
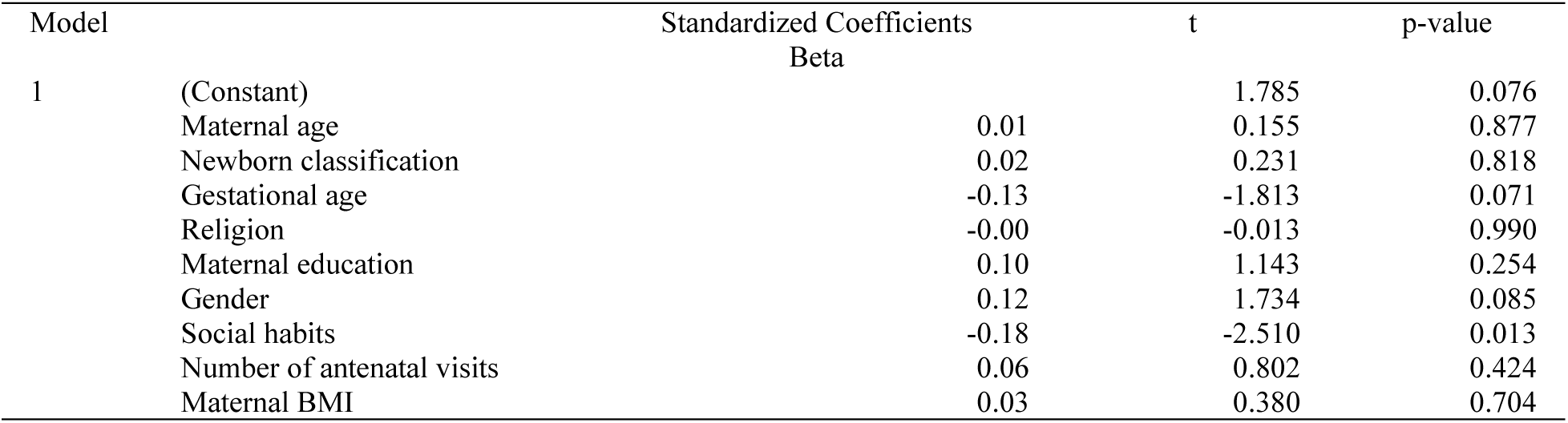
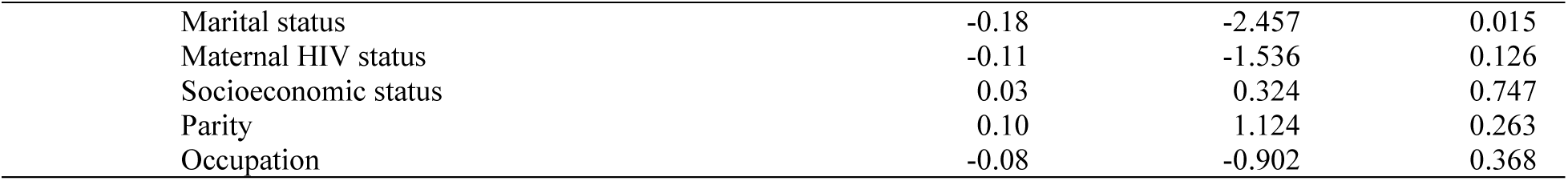
Multivariate linear regression analysis of maternal and neonatal characteristics associated with time-interval to breastfeeding initiation among babies delivered by cesarean section, n=205

### Significant characteristics of mothers and newborns in the cesarean section group with delayed breastfeeding initiation

Overall, the magnitude of delayed initiation of breastfeeding and prelacteal feeding, among the 529 babies were 17.0% (90) and 6.0% (32), respectively. Delayed breastfeeding initiation and prelacteal feeding rates among the 205 babies delivered by c-section were 37.6% (77) and 12.7% (26), respectively. The rate of prelacteal feeding among the c-section babies, who experienced delayed breastfeeding initiation was 46.2% (12 out of 26). The female newborns were less likely to experience delayed breastfeeding initiation compared to their male counterparts, p=0.073; OR= 0.59 (95% CI 0.33, 1.05). The babies that received prelacteal feeding were more likely to have experienced delayed breastfeeding initiation p=0.333; OR= 1.49 (95% CI 0.66, 3.45), compared to those, who were not fed prelacteal feeds. Mothers, who were exposed to potentially harmful substances during pregnancy were more likely to delay in breastfeeding initiation, p= 0.001; OR 3.14 (95%CI 1.56, 6.31). In Table 4, the Mann-Whitney U test (non-parametric independent T-test) analysis of the median breastfeeding initiation time-interval between the married and unmarried showed no significant difference (p=0.249). Breastfeeding initiation interval was 1 hour in both groups.

**Table 4:** Crosstabulation of significant factors and delayed breastfeeding initiation among babies delivered by cesarean section, n=205

| Newborn factors | p-value | Breastfeeding initiation |  | Total (100.0) |
| --- | --- | --- | --- | --- |
|  |  | Delayed (%) | Timely (%) |  |
| **Sex** | 0.073; OR 0.59 (95% CI 0.33, 1.05) |  |  |  |
| Female |  | 28(30.8) | 63(69.2) | 91 |
| Male |  | 49(43.0) | 65 (57.0) | 114 |
| **Prelacteal feeding** | 0.333; OR 1.49 (95% CI 0.66, 3.45) |  |  |  |
| Yes |  | 12 (46.2) | 14 (53.8) | 26 |
| No |  | 65 (36.3) | 114 (63.7) | 179 |
| **Maternal Factors** |  |  |  |  |
| **Marital status** | 0.206; OR 0.45 (95% CI 0.14, 1.41) |  |  |  |
| Unmarried |  | 4 (22.2) | 14 (77.8) | 18 |
| Married |  | 73 (39.0) | 114 (61.0) | 187 |
| **Exposure to social habits (smokes, alcohol, herbs)** | 0.001; OR 3.14 (95% CI 1.56, 6.31) |  |  |  |
| Yes |  | 25 (59.5) | 17 (40.5) | 42 |
| No |  | 52 (31.9) | 111 (68.1) | 163 |
| **Total** |  | **77 (37.6)** | **128 (62.4)** | **205(100.0)** |

Vaginal delivery was less likely to result in delayed initiation of breastfeeding (p<0.000; OR=0.18; 95% CI 0.07, 0.41) compared to cesarean section delivery. The median time interval from birth to breastfeeding initiation was 1 hour regardless of mode of delivery, however, the interquartile ranges for those in the cesarean section delivery group was significantly wider (IQR 1, 2 hours) than for those in the vaginal delivery group (IQR 1, 1 hour), p<0.000.

## Discussion

This study examined the burden and determinants of delayed breastfeeding initiation among mothers, who delivered by c-section in a rural district hospital in Rwanda, East Africa. The overall burden of delayed breastfeeding initiation was 17.0%; among mothers, who delivered by cesarean section, it was 37.6%; and among mothers that delivered vaginally, it was 4.0%, p<0.001. Prelacteal feeds were given to 12.7% of babies that were delivered by cesarean section, compared to 1.9% delivered vaginally, p<0.001, OR 1.47. These rates were higher than what were observed at rural and national levels in the same country by WHO.^48^ The preacteal feeds included plain water, glucose water, herbs or honey, which may be harmful to the newborn. Prelacteal feeding and delayed initiation of breastfeeding are strong risk factors for late onset neonatal sepsis.

The small and sick babies, who were likely to be unable to commence breastfeeding/breastmilk feeding immediately following birth^49^ and thereby increase further, the rate of delayed breastfeeding initiation, had already been excluded from the study. However, despite this precaution, prematurity still remained a deterrent to timely breastfeeding initiation as 11 out of the 26 (42.3%) healthy preterm babies in the cohort of 529 babies in this study, experienced delayed breastfeeding initiation.^49^ The ‘nutritive sucking pathway’ development, coordination of the suction-deglutition-respiration cycle and the breast-seeking reflex in the preterm baby are largely immature and contribute to this delayed breastfeeding initiation. ^49^

In the general population, Rwanda appears to be doing well in terms of timely initiation of breastfeeding, however, this improvement does not appear to extend to babies born via cesarean section delivery in rural areas. A higher percentage (37.6%) of the mothers, who delivered by cesarean section, delayed in initiating breastfeeding, compared to those (4.0%), who delivered vaginally, p<0.001. Though this Rwanda rate (37.6%) was lower than the global 60.3% delayed breastfeeding initiation rate, as reported by WHO;^12^ it was higher than the 31.4% observed at Mulago hospital, a national tertiary teaching health facility in Uganda.^50^ WHO, considers this rate of 37.6% as ‘good’ as it falls within the range of 11-50% on the infant and young child feeding assessment tool.^51^

Household water consumption was strongly related to both cesarean section delivery and delayed breastfeeding initiation. In this study, 94.4% or 37.7% of mothers, who delayed in initiating breastfeeding or who delivered by c-section, respectively, utilized water fetched from household sources.^52^ Unexpectedly, mothers, who used water sources in their home yards, were more prone to deliver by cesarean section, than those who had to go and fetch water from outside their homes, usually from the USAID and Water-for-life water bore holes, sunk in strategic places in the village. As earlier mentioned in this paper, the stored possibly contaminated water (with pathogens and toxins) may be a source of recurrent infections, that may have led to complications during pregnancy (high risk pregnancies) and hence, c-section delivery.^52^

Similar to Appaih’s [2021] report on maternal and child factors associated with timely initiation of breastfeeding in sub-Saharan Africa, married mothers were less likely to initiate breastfeeding within 1 hour after birth, compared to unmarried mothers. In this study, 39.0% of married mothers, who delivered by c-section, compared to 22.2% of unmarried mothers, delayed in initiating breastfeeding. This difference in the 2 groups was, however, not significant. This could be as a result of cultural influence from parents and in-laws.

Of note, prenatal exposure to potentially harmful social habits (such as tobacco, alcohol, herbs) contributed significantly to both c-section delivery and delayed breastfeeding initiation, as 42 (63.6%) of the 66 mothers, who were exposed, delivered by c-section.

The magnitude of cesarean section delivery was 38.8% in this study, and this rate could have been higher, if the deliveries of the sick and very small babies, that were excluded from the study, were considered. This c-section rate was much higher than the WHO safety limit of 10-15%, and indeed higher than the rates documented in several countries in sSA. Cesarean section delivery was a major hinderance to timely breastfeeding initiation in the study.

Kalisa [2015] explained the delay in breastfeeding initiation following c-section delivery to be as a result of maternal exhaustion, sleepiness and severe abdominal pain.^50^ Effects of anaesthesia delaying the onset of lactation or associated neonatal respiratory distress may further compromise timely initiation of breastfeeding in babies delivered by c-section.^26^

Frequency of antenatal care visits was generally poor in the study, as only 43.1% of mothers attended ANC ≥4 times during pregnancy. And it appeared that the majority of those, who had high frequent visits (>6), had complicated pregnancies, that may have led to c-section deliveries. This is evidenced by the fact that 80% of the mothers, who frequented the ANC >6 times, delivered by c-section, and 40% of these same mothers delayed in initiating breastfeeding. All the 3 out of 529 mothers (0.6%), who did not attend ANC initiated breastfeeding within 1 hour after birth and only 1 (33.3%) of them had a cesarean section delivery. Of note, access road to the hospital was hilly and very difficult to navigate, especially during the raining season (and it rains virtually every week in Rwanda), hence, the mothers appeared to seek hospital services, only when they have health problems.

In this current study, mothers, who were HIV positive, were more likely to undergo c-section delivery (88.9%, OR 7.14) than those who were HIV negative (36.1%). HIV-indicated cesarean deliveries, mainly to prevent MTCT peaked at 48% in 2004, then dropped to 12% by 2013 in 2 observational studies conducted in the USA, involving 6444 HIV positive women.^53^ On the contrary, in places such as Malawi,^54^ where a retrospective study involving 17 health facilities and 62,033 women, observed that c-section delivery (rate of 19.1%) was largely determined by maternal and fetal complications, rather than HIV positivity. In this Malawi study, HIV positive mothers were less likely to be delivered by c-section, possibly because of risk of occupational HIV transmission. ^54^

As expected, mothers with moderate parity and those that do not use charcoal for cooking and house warming were less likely to deliver by c-section in this current study. Weber et al. (2020) in Ghana Accra metropolis, reported that prenatal exposure to polluting cooking fuel such as wood/charcoal (33%) led to perinatal mortality and fetal low apgar score, among 1010 pregnancies in a prospective cohort study.^55^ Systematic review by Luo et al (2023) also linked use of solid fuel to SGA and LBW births.^56^ The mechanism involved could be the fetal cardiorespiratory effects from inflammation and oxidative stress induced by reactive oxygen and nitrogen species generated by inhaled pollutants. These are fetal complications that could have necessitated c-section deliveries.

For non-medical causes, maternal age and occupation were significant determinants of c-section delivery in this study. Majority of the teenage (56.5%) and entrepreneur (76.9%) mothers, mothers who were heavier and/or shorter were likely to undergo cesarean section delivery in this current study. Fetal size did not play a dominant role in the decision to deliver by c-section in the multivariate analysis of this study.

Maternal socio-economic status was not a significant determinant of mode of delivery in this study, contrary to the claims by WHO (2017), Betran et al (2016) and Bayou et al (2016), who documented that socioeconomic status was a significant determinant of c-section delivery in LMICs. However, only 17 (3.2%) of the mothers in this study came from rich homes and this may have affected the logistic regression analysis. Though, maternal socioeconomic status and educational level did not significantly affect the mode of delivery, the rich, highly educated and entrepreneurs were more likely to deliver by c-section compared to their poor, lowly educated and low skilled counterparts. This was similar to Arunda et al. (2020)’s findings, where the odds of c-section delivery was higher amongst mothers from richest households, highly educated, and managers, compared to middle class, un-insured, poorly educated and unemployed mothers ^57^.

Where, timely breastfeeding initiation may not be possible, because of maternal complications, post-cesarean section delivery and when breastfeeding is desired, in some instances, antenatal breastmilk expression (aBME), human donor milk from breastmilk bank, screened surrogate or wet nursing, if available, may be practiced until mother is able to breastfeed. The last resort in these situations should be the offer of breastmilk substitutes such as infant formula, only when breastmilk is completely unavailable. Due to the lack of safe water in this village, infant formula will be an unlikely solution to this problem for this community, as this would compromise feeds preparation and result in diarrhea diseases. Moreover, the prohibitive cost of infant formula is out of the reach of most families in this rural population. MoH Rwanda, discourages the use of infant formulas.^29^ Human Donor milk banks are rare in sub-Saharan Africa. Screening of potential wet nurses for transmissible diseases, may not be feasible, if they do not consent, and also if, the appropriate laboratory test is unavailable in the rural laboratories, hence, the risk of infecting the newborn may be increased in these situations. Otherwise, babies born by cesarean section should be closely monitored for timely initiation of breastfeeding to discourage prelacteal feeding.

The interventions available in resource limited settings to alleviate the problems of delayed initiation of breastfeeding among mothers, especially, those who deliver by c-section, could include provision of high calorie, high protein porridge with supplements to ensure adequate hydration for mothers in the post-natal ward; pillows for more comfortable breastfeeding positioning; screens for mothers to breastfeed or express breastmilk privately; provision of refrigerator and materials for hospital storage of expressed breastmilk, and educational posters promoting early initiation of exclusive breastfeeding and Kangaroo Mother Care (KMC). Training of health care workers and expert mothers (serving as peer counsellors) in infant feeding; creating breastfeeding-friendly, caring environment for mothers with a focus on one-to-one as well as group counselling [48]. Antenatal breastmilk expression (aBME) also appears to be promising, aimed at collection of colostrum for post-partum use, in anticipated situations, where mothers may not be able to initiate breastfeeding in time. aBME involves expressing breast milk from 32 to 37 weeks gestation to the time of delivery, about once or twice per day, each session lasting between 5 to 10minutes. Manner of expression of breastmilk may be by nipple massage, nipple rolling or breast massage. Antenatal breastmilk expression is possible because colostrum production begins right from mid-pregnancy, around 12-18weeks gestation. aBME has been shown to hasten lactogenesis II, decrease postpartum breast engorgement, avoid the need for breastmilk substitutes (infant formula), improve blood glucose stabilization in newborns at risk of hypoglycemia, and increase exclusive breastfeeding maintenance up to 6 months postpartum. However, antenatal breast stimulation leading to oxytocin release have been shown to induce of preterm labour or miscarriage,^58^ and these concerns have slowed the adoption of this approach. Effectiveness, acceptability and safety of aBME needs to be further explored in resource-limited countries in future researches.

## Conclusion and Recommendations

From the observations made in this study, HIV positivity and non-medical causes still drive the over-use of cesarean section for delivery in the rural areas, resulting in unnecessary surgeries, with their associated risks. This practice is also a hindrance to timely breastfeeding initiation and it is promoting prelacteal feeding in a magnitude that requires intensification of pro-breastfeeding interventions such as BFHI and Early essential newborn care, to quickly curb the rising trend and protect newborn survival and wellbeing. The use of probably unsafe water that was linked to increased c-section deliveries and delayed breastfeeding initiation in the village, require the water supplies and safety to be further investigated and improved. Prenatal exposures to harmful substances should be addressed during antenatal counselling. Rural antenatal care visitation, use of charcoal as cooking fuel, health facility delivery and breastfeeding practices need to be closely monitored and regulated. Policies and protocols need to be strengthened to guide the use of cesarean section for deliveries and improve BFHI practices amongst health workers in rural areas. Special health promotion (including maternal education), intervention (including antenatal breastmilk expression) and healthcare provider support to facilitate improved antenatal care acceptance, immediate or early skin to skin contact, and early initiation of breastfeeding for post-operative patient should be considered.

## Data Availability

All data produced in the present study are available upon reasonable request to the authors

https://www.example.com

## Author Contributions

The corresponding author (Dr Adenike Oluwakemi Ogah) conceived and designed the study, collected data and conducted data analysis, interpreted the results, and drafted the manuscript. Dr Monica Kapasa reviewed and edited the manuscript for intellectual content. The authors approved the final manuscript for submission.

## Acknowledgements

The author is extremely grateful to the participants involved in this study, to the staff of Gitwe Hospital and clinic in Rwanda and to the research team. I am also indebted to my supervisors (Prof Andre Venter and Prof Corina Walsh), and my statistician Prof Gina Joubert, who analysed all the data for my PhD thesis.

